# COVID-19 DYNAMICS: A HETEROGENEOUS MODEL

**DOI:** 10.1101/2020.05.04.20090688

**Authors:** Andrey Gerasimov, Georgy Lebedev, Mikhail Lebedev, Irina Semenycheva

**Affiliations:** I.M. Sechenov First Moscow State Medical University, 2-4 Bolshaya Pirogovskaya Street, 119991 Moscow, Russia; Federal Research Institute for Health Organization and Informatics, 11 Dobrolubova Street, 127254, Moscow, Russia; Center for Bioelectric Interfaces, Institute of Cognitive Neuroscience, National Research University Higher School of Economics, 13 Myasnitskaya Street, Building 4, 101000, Moscow, Russia

## Abstract

The ongoing Coronavirus disease 2019 (COVID-19) epidemic is different from the previous epidemic of severe acute respiratory syndrome (SARS), which demands a rigorous analysis for the selection of anti-epidemic measures and their lifting when the epidemic subsides. Here we estimate the basic reproductive number for COVID-19 and propose a dynamical model for the time course of infection number. With this model, we assessed the effects of different measures for infection risk control. The model is different from the previous ones as it models the population as heterogeneous, with subpopulations having different infection risks. Our analyses showed that after this heterogeneity is incorporated in the model, several characteristics of the epidemic are estimated more accurately: the total number of cases and peak number of cases are lower compared to the homogeneous case, the early-stage growth rate in the number of infection cases is little affected, and the decrease in the number of infections slows down during the epidemic late stage. The comparison of our model results with the available data for COVID-19 indicates that the anti-epidemic measures undertaken in China and the rest of the world managed to decrease the basic reproductive number but did not assure an accumulation of sufficient collective immunity. Thus, the epidemic has a high likelihood to restart, which necessitates a careful approach to lifting the quarantine measures.

## Introduction

The new coronavirus infection subsequently named Coronavirus disease 2019 (COVID-19) was first identified in the city of Wuhan. The initial cases of COVID-19 were reported in late November 1919 (Zhu, Xie et al. 2020). A month and a half after the first reports, on January 15, there were only 41 cases on the record. Then, the number of cases grew rapidly (Boldog, Tekeli et al. 2020, Liu, Magal et al. 2020, Zheng, Xu et al. 2020, Zhou, Yu et al. 2020). The number of cases increased by more than one thousand from January 15 to February 15. Starting from January 2020, China took extreme quarantine measures. In mainland China, incidences of the disease started to decline, but both the number of countries with reported cases and the incidence rate kept increasing (Arab-Mazar, Sah et al. 2020, Heymann and Shindo 2020).

COVID-19 is caused by the virus SARS-CoV-2. Clinical manifestations of the disease resemble those of severe acute respiratory syndrome (SARS). The mortality rate is lower than in SARS (Zhou, Yu et al. 2020) but the incidence rate is significantly higher. The current COVID-19 epidemic differs in several aspects from the previous one caused by SARS, which was finally extinguished (Cossarizza, De Biasi et al. 2020, Kucharski, Russell et al. 2020, Kuniya 2020, Leung 2020, Mizumoto, Kagaya et al. 2020, Parodi and Liu 2020). First, COVID-19 has a higher basic reproductive number, R_0_, than SARS. Second, in contrast to SARS, causative pathogen transmission in COVID-19 starts before the end of the incubation stage of the disease. Third, unlike SARS, many (and possibly most) cases of COVID-19 are asymptomatic, but they are accompanied by a spread of causative pathogen. These features of COVID-19 lower the optimism for the prospect that the current epidemic will be successfully controlled.

At the time of this writing, several issues remain unclear regarding the spreading of this pathogen around the globe, the ways to avoid mass morbidity, estimation of the total incidence rate, and the risk that the incidence rate could start growing after the emergency anti-epidemic measures are partially cancelled.

Here we simulated COVID-19 dynamics using a model that accounted for the heterogeneous composition of the population with subgroups affected differently by the disease. With this heterogeneous model, we assessed the role of anti-epidemic measures and changes in the number of infected and recovered individuals. Our results explain the available data on the epidemic and predict its future development after the anti-epidemic restrictions are lifted.

## Methods

Our dynamical model accounts for the heterogeneity of infection risk across different age groups. This is important because the risk of developing COVID-19 strongly depends on patient age (Lee, Hu et al. 2020, Liu, Chen et al. 2020, Ruan, Yang et al. 2020) and because measures against the disease spread include isolation of the elderly individuals. Given these factors, it is important that population heterogeneity by the infection risk, α, is incorporated in the model of disease dynamics.

In our model, I(α,t) and S(α,t) are the proportions of infected and susceptible people, respectively, α is infection risk, t is time, and dF(α) is the across-population statistical distribution of the infection risk. (∫ *dF* (*α*) = 1.) For an infinite isolated population, epidemic dynamics is defined by the set of differential equations (Герасимов and Разжевайкин 2008):

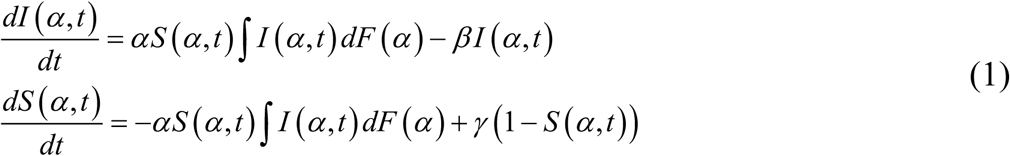

where 1/β is the average disease duration from the infection till the end of pathogen transmission, and 1/γ is the average lifespan for people with lifelong immunity or the average duration of sustained immunity for the people with transient immunity. The relationship between infection risk, α, and the basic reproductive number is given by the equation:

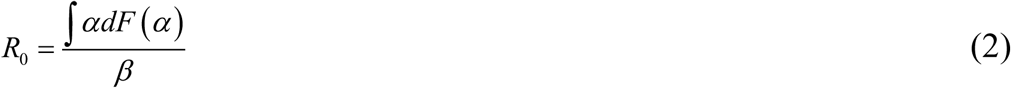

If an epidemic is considered that continues for several months, then we can neglect the term *γ* (1 − *S* (*α*, *t*)) that defines population renewal. In this case, the dynamical equations can be rewritten as:

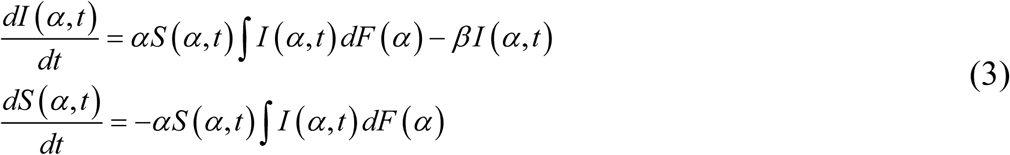

The disease progression is usually described as discrete daily samples, where the variations of people activities throughout the day are averaged out. Accordingly, if J(α,k) is the portion of infected people on day k then equation (3) can be rewritten to have discrete steps:

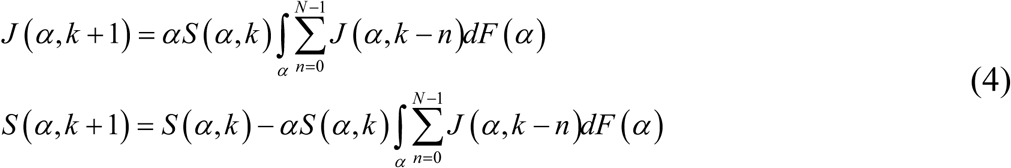

where N is disease duration in days from the infection onset till the cessation of pathogen transmission, and *R*_0_ *= N ∫ αdF* (*α*).

The equations 1, 3 and 4 belong to the susceptible–infected–recovered (SIR) class of models of an epidemic process. As followers from the model name, population members can be in one of three states: susceptible, infected, and immune. The susceptible-exposed-infectious-recovered (SEIR) models describe the initial period of an epidemic more accurately. In these models, an additional state is added, exposed (infected but sterile), that corresponds to the very start of an infection. In this model, the following equation describe the epidemic dynamics:

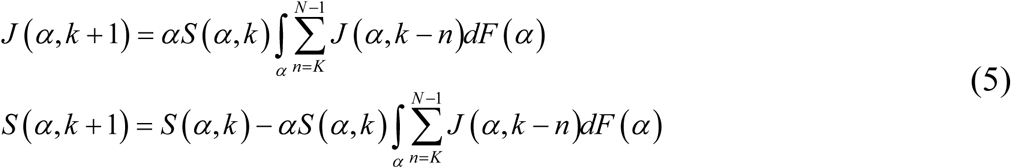

where K is the duration of sterile period in days, and *R*_0_ *=* (*N* − *K*)∫*αdF* (*α*).

The model that we implemented for COVID-19 contained several additional assumptions. First, we assumed that the disease incidence initially increases exponentially in a nonimmune population. The rate of growth depends on the reproduction number, R_0_, and the disease duration – the factors that determine the distribution of time intervals between sequential infections. The lower limit for the time interval between infections is the time from the infection onset till the beginning of virus shedding, and the upper limit is the time from the infection onset till the end of shedding plus the duration of pathogen preservation in the external environment.

According to the available data, the growth rate is significantly higher for SARS-CoV-2 than for SARS when comparable durations of the disease are considered. Indeed, at the initial stage of SARS epidemic in 2003, the number of cases tripled during the month of April from 2,000 to 6,000 (Chan-Yeung and Xu 2003, Deng and Peng 2020). By contrast, in the second half of January 2020, the number of cases in Wuhan tripled in 3-4 days (i.e. a 40% increase per day).

Quantifying the time interval between infections is difficult even for the well-studied infectious diseases. This is because the beginning of the causative pathogen shedding does not always coincide with the end of incubation period. Additionally, the time interval between infections is affected by such factors as changes in the intensity of the causative pathogen shedding at different stages of the disease, changes in patient behaviour, and person-to-person variability. In particular, it is necessary to consider all forms of infection including healthy carrier states.

Given these considerations, we based our model on a simplified assumption that during the entire infectious period the infection rate remains constant, and the duration of infectious period, t_1_, is equal to the duration of sterile period, t_2:_ t=t_1_= t_2_. We performed modelling for different values of t.

## Results

We used our dynamical model to assess several factors that could slow down the daily growth of the incidence rate: a sufficiently high cumulative incidence, growing collective immunity in high-risk groups, and anti-epidemic measures.

Table 1 shows how daily growth in the number of infection cases depends on R_0_ and t. The estimation of R_0_ based on the daily growth is only approximate because of the imprecise estimation of the sterile and infectious periods and because virus shedding by an infected person and his/her interactions with the other people change in time. Additionally, infection control measures result in a decrease in the number of people interacting with the infected person and, consequently, the daily growth in the number of cases. For example, the daily growth was 25% in Moscow in the beginning of COVID-19 epidemics, and after the introduction of quarantine measures it decreased to 15%.

**Table 1.** Daily increase in the number of infection cases at the epidemic initial stage as the function of reproduction number, R_0_, and the duration of sterile/infectious period, t.

| $R_0$ | $t$, days | | | |
| --- | --- | --- | --- | --- |
|  | 3 | 4 | 5 | 6 |
| 2 | 19,2% | 17,1% | 15,3% | 13,9% |
| 3 | 32,5% | 28,8% | 25,9% | 23,5% |
| 4 | 42,9% | 38,0% | 34,2% | 31,1% |
| 5 | 51,7% | 45,8% | 41,2% | 37,5% |
| 6 | 59,3% | 52,6% | 47,3% | 43,1% |

Figure 1 shows the number of infected people as a function of time for a city with 10 million inhabitants; t is set to 5 days, and R_0_ is set to 2 or 4. Here the results of a homogeneous model (solid lines) are compared with the results of a heterogeneous model (dashed lines). In the heterogeneous model, infection risk has a uniform distribution between 0 and 2R_0_. It is evident from this analysis that the overall incidence is lower when the heterogeneity factor is incorporated in the model. A noticeable slowdown in the incidence rate, however, begins to manifest itself only when the overall incidence has reached a sufficiently high value. In this example, we are considering a rather distinct heterogeneity, whereas heterogeneity is much less expressed in the case of usual airborne infections, which have high infection risk for everybody.

**Figure 1.**
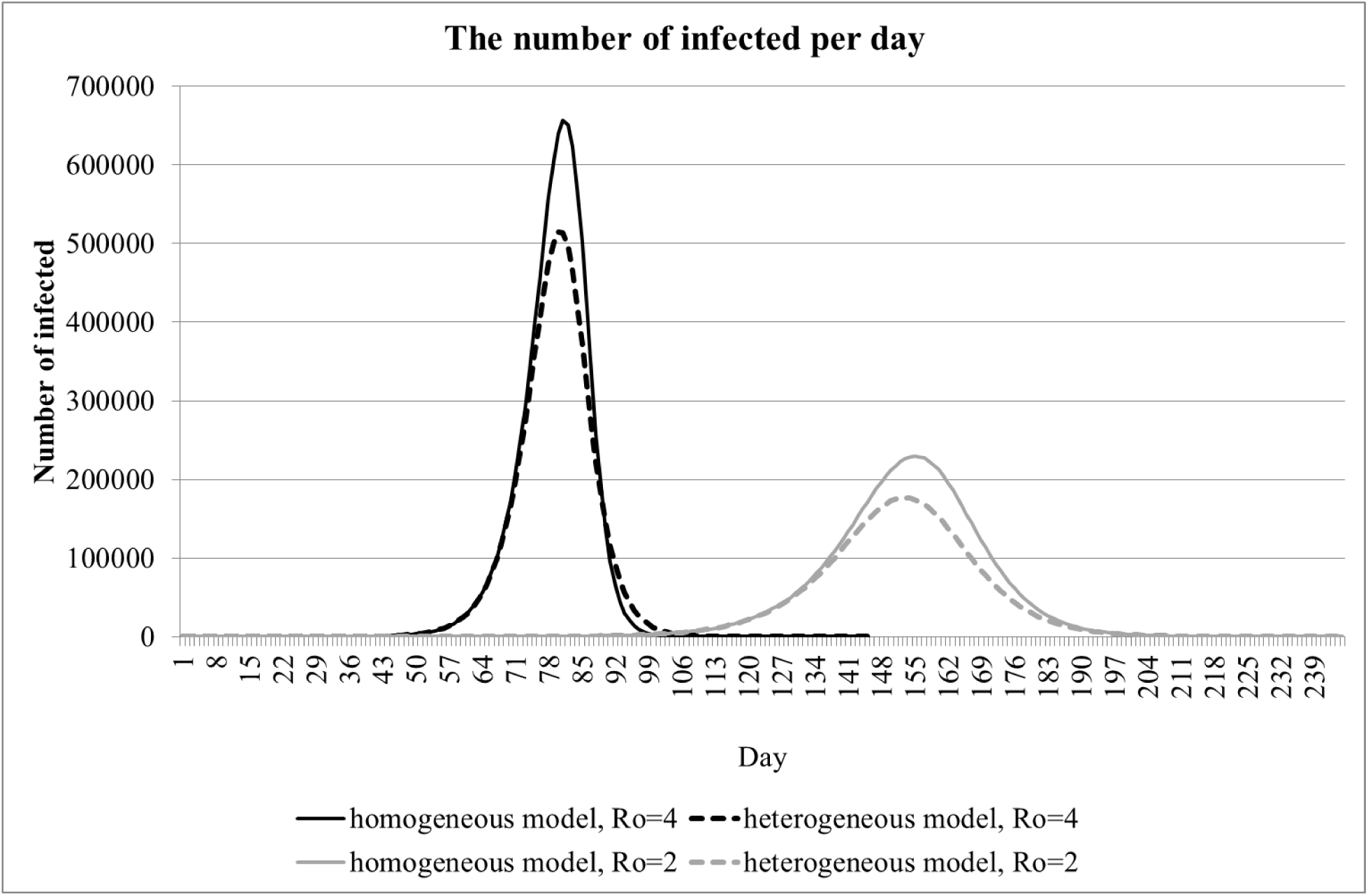
The dynamics of the number of infections per day in a 10-million population. The curves for homogeneous (solid lines) and heterogeneous (dashed lines) are shown for R_0_ equal to 2 and 4.

Anti-epidemic measures strive to reduce the COVID-19 infection rate even before it starts to decrease because a substantial portion of the population (including hidden cases) is affected. We modeled the effect of anti-epidemic measures by decreasing R_0_ from 4 to 2 (Figure 1). With these settings, the modeled anti-epidemic measures of moderate intensity shifted peak incidence forward in time and reduced the peak value. However, the total incidence does not change appreciably, as evident from the widening of the curve.

For the selection of appropriate anti-epidemic measures, it is important to measure R_0_ early in the epidemic. One can estimate R_0_ based on the disease duration and the growth of incidence rate in the beginning of an epidemic, when the growth is exponential (Table 1). At the exponential-growth stage, daily increase in the total number of cases, expressed as percentile, is constant. This metric is calculated as the ratio of the number of cases on a given day to the sum of cases during the preceding days. Figure 2 shows the dynamics of this ratio for several regions, including China provinces and the other countries. Points are median values, and error bars on the curve for the China provinces (red line) are quartiles. The value of 100% corresponds to the number infected people doubling on a given day.

**Figure 2.**
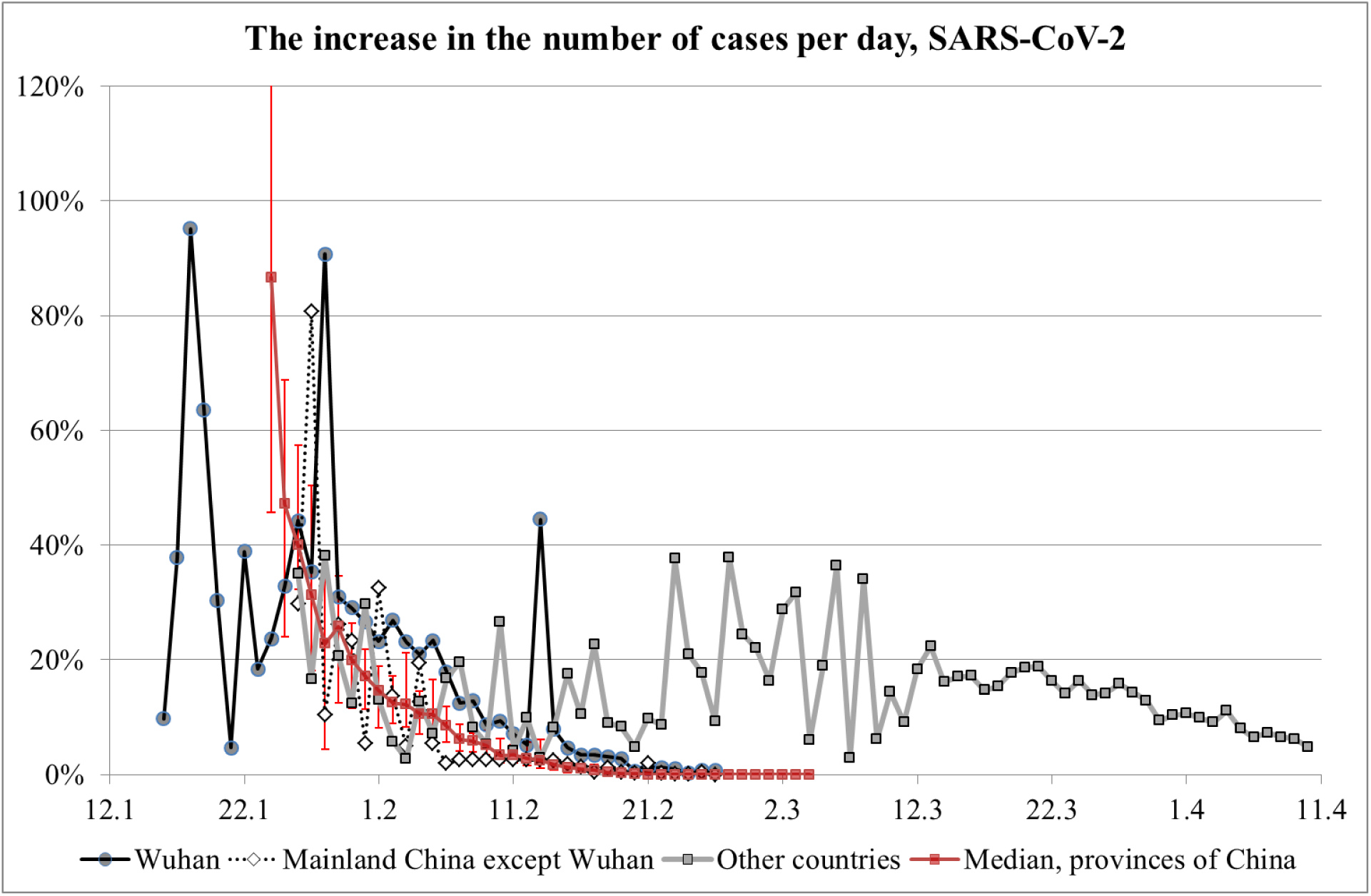
Incidence rate growth rate in different regions.

For Wuhan data, the early 90% peak in growth rate can be disregarded because it corresponds to the very beginning of the disease diagnostics and low samples. Next, when the diagnostics was ongoing, the growth rate peaked to 40%, which corresponds to R_0_ of 2.5 (see Table 1), then decreased to 20% (i.e., R_0_ of 4) in early February and clearly terminated in mid-February. Given the relatively low overall number of infections, this marked slow-down of infection progression occurred because of the anti-epidemic measures, not because of an accumulation of collective immunity. A note should be made about the 45% surge in growth rate in Wuhan on February 13. It is related to a change in the methodology for calculating the number of cases. On that day, the cases previously considered as questionable were added to the report. Thus, the graph for Wuhan matches the prediction of our model where R_0_ and the number of infections decrease because of anti-epidemic measures. A similar dynamic is seen for the rest of the world, where anti-epidemic measures were also undertaken and resulted in the growth rate decrease after March 13.

## Discussion

In this study, we used a heterogeneous model to simulate COVID-19 epidemic, and obtained more accurate results compared to the simpler, homogeneus models. Heterogeneity is an important factor for most infectious diseases. For example, for COVID-19, there is a population of individuals who are infected but do not show noticeable symptoms (Bai, Yao et al. 2020, Mizumoto, Kagaya et al. 2020). These individuals could be omitted from the medical reports. These people would transmit the infection to others and obtain specific immunity at the end of their infection period. These cases could be underreported because polymerase chain reaction (PCR), the existing methodology for diagnostics, cannot detect the individuals recovered from the disease. Additionally, there is a bias toward testing mostly the patients who have clinical symptoms. Furthermore, there is an age-related heterogeneity as the disease incidence increase with age (Liu, Chen et al. 2020). The average age of patients with clinical symptoms is over 50 years old whereas there are virtually no reported cases of infected children – the distribution that is at odds with the typical risk of infection for airborne infections, which is high for all age groups.

Our model accounts for heterogeneity of infection risk and provides more realistic estimates of the number of infections needed to accumulate for the epidemic to slow down. The model also allows to assess the effect of anti-epidemic measures. Accordingly, we looked at two factors that contributed to the lowering of the morbidity growth rate in China and other countries: (1) the anti-epidemic measures, and (2) the accumulation of a sufficiently large proportion of the subpopulation that includes both the infected cases and cases of recovery from the illness in any form, including a mild form without pronounced clinical symptoms. We modeled the first factor by decreasing R_0_ and found a slow-down of the epidemic progression while the total number of eventually infected people remained unchanged. Thus, the second factor is quite important according to our model because it explains the conditions that guarantee that an infection has ended and would not restart.

The comparison of our model results with the data for Wuhan and the rest of the world indicates an R_0_ of 4 at the epidemic starts, followed by a decrease to 2.5 after the introduction of anti-epidemic measures and a cessation of epidemic growth when the measures become strict. Yet, the total number of infected people is relatively low at this point, which could be insufficient for the second factor to guarantee that the epidemic has completed. Note that the decrease in incidence growth is almost the same for all Chinese provinces regardless of huge discrepancies in morbidity levels among the provinces. For example, by March 4 the number of recorded cases in Hubei Province reached 67,466 while the median number of cases in other 35 provinces amounted to just 245 cases. Such dynamic is consistent with anti-epidemic measures taking their effect. Indeed, if the decrease in the growth rate were due to an accumulation of unreceptive cases, the decrease rate in the other provinces would have varied greatly. Moreover, a similar dynamic occurred for the rest of the world.

Based on these results, we draw the following conclusions:

1. The characteristics of COVID-19 differ markedly from SARS, which makes it hard to contain the disease spread to an affected territory unless the anti-epidemic measures are strict.
2. In the absence of effective anti-epidemic measures, more than 1% of the population could get infected. Should this happen, most cases will occur during the period of several months, which will cause great problems to the treatment of patients.
3. After lifting the emergency quarantine measures, the epidemic could restart because of an insufficient collective immunity level. This course of events should be seriously considered when “reopening” provinces and countries.

## Data Availability

The manuscript reports a mathematical model that can be applied to open-access data: https://datascience.nih.gov/covid-19-open-access-resources

https://www.cdc.gov/library/researchguides/2019novelcoronavirus/databasesjournals.html

